# Development and acceptability of a support intervention for families after sudden cardiac death in the young

**DOI:** 10.1101/2023.10.16.23297063

**Authors:** Laura Yeates, Amy Baker, Karen Gardner, Natalie Stewart, Laura Catto, Judy Do, Sue Timbs, Felicity Leslie, Christopher Semsarian, Belinda Gray, Leesa Adlard, Jodie Ingles

**Affiliations:** Genomics and Inherited Disease Program, Garvan Institute of Medical Research, and UNSW Sydney, Sydney, Australia; Faculty of Medicine and Health, The University of Sydney, Sydney, Australia; Department of Cardiology, Royal Prince Alfred Hospital, Sydney, Australia; Academic Unit of General Practice, Australian National University, Canberra, Australia; End unexplained cardiac death (EndUCD.org); Agnes Ginges Centre for Molecular Cardiology at Centenary Institute, The University of Sydney, Sydney, Australia

## Abstract

Sudden cardiac death (SCD) in the young (<35years) can be due to an inherited cardiovascular condition. The impact of SCD on the surviving family is significant, with high rates of symptoms of posttraumatic stress and prolonged grief. Using stakeholder codesign we developed COPE-SCD: an online community supporting families after SCD. The intervention includes a website and four online support sessions (general information on SCD, navigating uncertainty, coping with grief and loss both individually and as a family). Here we aim to develop content and assess the acceptability of the COPE-SCD intervention. Participants were recruited from the Genetic Heart Disease Clinic, Royal Prince Alfred Hospital, Sydney, Australia and EndUCD.org, a patient organisation. Demographic and psychological measures were collected at baseline. ‘Think aloud’ interviews were conducted to assess the website. Online sessions were assessed with post session questionnaires and qualitative interviews. Both interview schedules and questionnaires were mapped to seven constructs of the Theoretical Framework of Acceptability. Website and online session content were developed. Six ‘think aloud’ interviews were conducted to assess the website, including feedback on content and layout. Twelve participants, in two groups, completed the four online sessions. Overall, participants liked both parts of the COPE-SCD intervention, particularly the opportunity for peer support. They found the intervention acceptable when considering the seven constructs of the theoretical framework of acceptability. Further work is needed to assess the effectiveness of the intervention as it’s implemented into clinical practice.

**What is known about this topic:** Sudden cardiac death due to an inherited cardiovascular condition has a devastating impact on the surviving family, with high rates of psychological distress. Previous research has shown more psychosocial support for family members is needed.

**What this paper adds to the topic:** We describe the development and acceptability of a support intervention for families after sudden cardiac death in the young. The intervention combined information and peer support and was found acceptable to individuals with a family history of sudden cardiac death.

## INTRODUCTION

Care of families after sudden cardiac death (SCD) of a young person often focuses on clinical screening of first-degree relatives and postmortem genetic testing. However, previous work has highlighted the psychological impact of SCD on the surviving relatives (Ingles et al., 2016; McDonald et al., 2020; Steffen et al., 2020; Yeates et al., 2013). Indeed, the importance of psychological support has been recognised in recent guidelines and highlighted as a research priority (Marijon et al., 2023; Stiles et al., 2021). As a key member of the multidisciplinary team caring for these families, the genetic counselor is a ‘frontline’ source of care connecting families with further support as needed (Caleshu et al., 2016). Psychological support can include professional care from clinical psychologists and grief counselors, and there is increasing evidence for the benefits of peer support (Bartone et al., 2019; Steffen et al., 2020).

The experience of a young SCD brings unique challenges. Family members are dealing with the unexpected nature of the death, but also the possibility of a heritable cause that places other family members, including themselves, at risk of similar events (Stiles et al., 2021). Furthermore, for 40% of cases aged between 1-35 years, no cause of death is identified even after comprehensive postmortem investigation (Bagnall et al., 2016), leaving uncertainty as to why their family member died (Ingles & James, 2017). While general grief support groups exist and can be helpful in this setting, previous work has highlighted a want, and need, for among SCD families, to meet others with a similar experience(Steffen et al., 2020; van den Heuvel et al., 2024).

Following codesign focus groups (Yeates et al., 2022), COPE-SCD: an online community supporting families after SCD in the young was developed. The intervention has two parts: a website and a series of four online sessions. The website provides family members affected by SCD, their extended family and friends, and healthcare professionals with information about causes of SCD, clinical screening and genetic testing and coping with grief and loss. Recognising the internet is a widely utilised source of health information for Australians (Cheng & Dunn, 2015) a website was selected as a key part of COPE-SCD during the intervention’s codesign process (Yeates et al., 2022). Participants suggested a website could provide accurate, easily sharable, information to families, friends and healthcare professionals regardless of geographical location. It could include clear referral pathways and provide a platform for peer support. Although some websites exist internationally (*Cardiac Risk in the Young*, 2024; *Sudden Arrhythmia Death Syndromes (SADS) Foundation*, n.d.) information specific to the Australian context e.g. coronial processes and referral to clinical services, was requested (Yeates et al., 2022). The second part of the intervention involves online sessions: a series of four one-hour sessions to provide information and an opportunity for peer support. Internet-based peer support removes the need and cost to travel to attend a face-to-face group (van der Houwen et al., 2010) and allows more control over the level of engagement (e.g. turning camera off, taking a break, using a pseudonym). Internet based sessions were chosen for the COPE-SCD sessions in the codesign process, as they were a convenient forum to connect and interact with other SCD family members and access to experts for information (Yeates et al., 2022).

Implementation science has become necessary in medical research to effectively incorporate new health interventions into clinical care. Before considering implementation of a developed intervention, acceptability of the intervention must be assessed (Klaic et al., 2022). One widely used (Coffey et al., 2023; Paynter et al., 2023; Turnbull et al., 2023) framework to guide the researcher is the Theoretical Framework of Acceptability developed by Sekhon et al. (Sekhon et al., 2017). This framework outlines seven constructs of acceptability (affective attitude, burden, ethicality, intervention coherence, opportunity costs, perceived effectiveness and self-efficacy) to be considered when deciding whether an intervention is acceptable to participants and/or deliverers (Sekhon et al., 2017). Here we aimed to develop content for both the website and the online sessions and assess acceptability of the developed website via a ‘think aloud’ analysis and interview; and assess the acceptability of the online sessions using a mixed methods approach.

## METHODS

### Development of the website

Based on the topics raised in the codesign focus groups (Yeates et al., 2022), a website menu was drafted by LY with the research team. First draft of the content was primarily written by LY with members of the research team contributing as required based on their clinical expertise (e.g. clinical psychologist). Other experts in the team (cardiologists BG/CS and genetic counsellors) then reviewed pages relevant to their expertise. Once drafted, all content was reviewed by other genetic counsellors in the team (AB, NS, LC) to ensure consistency of language. The Flesch-Kincaid Grade level score in Microsoft Word was used to assess readability. Pages were also reviewed using the health literacy editor (https://shell.techlab.works/) which helps identify complex words and sentences within text (Ayre et al., 2023).

The website included an animated video. The video aimed to give a general overview of SCD and repeat key information that would often be given in the first phone call with a family from a genetic heart disease clinic. The video was designed to be easily shared with extended family and friends of the deceased person. Key points of the script were developed by the research team whilst working with the professional animator (www.animateyour.science).

### Development of the online sessions

Based on feedback from the codesign focus group (Yeates et al., 2022) the research team developed an outline for the online sessions. Genetic counselor (LY) and clinical psychologist (LA) further developed content based on the outline with content designed to complement the COPE-SCD website. Time was allocated in each session for participants to share their thoughts and experiences.

The online sessions comprised a set of four, one-hour sessions held over video conference, one week apart. They were facilitated by a genetic counselor and clinical psychologist, and a cardiologist attended when required. Each session had a different topic and focus, with the aim to provide information and opportunities for participant sharing and peer support. Session one focused on general introductions including an explanation of session format, housekeeping, confidentiality, and time for participants to introduce themselves and describe their experience. Session two aims to ensure participants gain an understanding of SCD and the need for family investigation. It covers the causes of SCD in young people, clinical screening of at-risk relatives and postmortem genetic testing. There is allocated time for a “question and answer” session with the team (including a cardiologist). Questions are also welcome throughout the other sessions. Session three focuses on grief and loss at an individual level, including how grief is experienced differently for different people, with an opportunity for sharing their personal journey. The final session covers grief and loss with a focus on managing variable coping styles within a family and their wider support network, including differences in grief reactions and the potential impact on family dynamics. In addition, the session includes discussion of how families approach anniversaries, birthdays and missed milestones.

### Participants

Ethics approval for this project was granted through Sydney Local Health District, RPA Zone, protocol number X21-0284. Participants were recruited in two ways: 1. From the Genetic Heart Disease Clinic, Royal Prince Alfred Hospital, Sydney, Australia and 2. Via End Unexplained Cardiac Death (EndUCD) organisation, a charity for Australians impacted by SCD (enducd.org). For the website review, participants were only recruited via the specialised genetic heart disease clinic, purposive sampling was used to obtain demographic diversity with respect to sex, age, and relationship to the deceased. Eight potential participants were invited by an experienced cardiac genetic counsellor (LY), who worked within this team. We included those aged ≥18 years, with a first-degree relative (including spouse) having experienced SCD.

For the online sessions, a purposive sampling approach was used to select families who had been seen at the clinic in the preceding six years (starting with most recent), and who had experienced the SCD of a relative. Efforts were made to recruit a range of demographics including relationship to the deceased. Where the death was recent (less than six months), families were not approached. Participants were invited to participate via phone and a follow-up email by a genetic counselor (LY). For EndUCD recruitment, the main contact for each family was identified by the EndUCD director (ST), who sent an email invitation and connected interested participants with the research team. Participants were ≥18 years and had self-reported English skills sufficient to complete a questionnaire and qualitative interview. A summary of the methods can be found in Figure 1.

**FIGURE 1.**
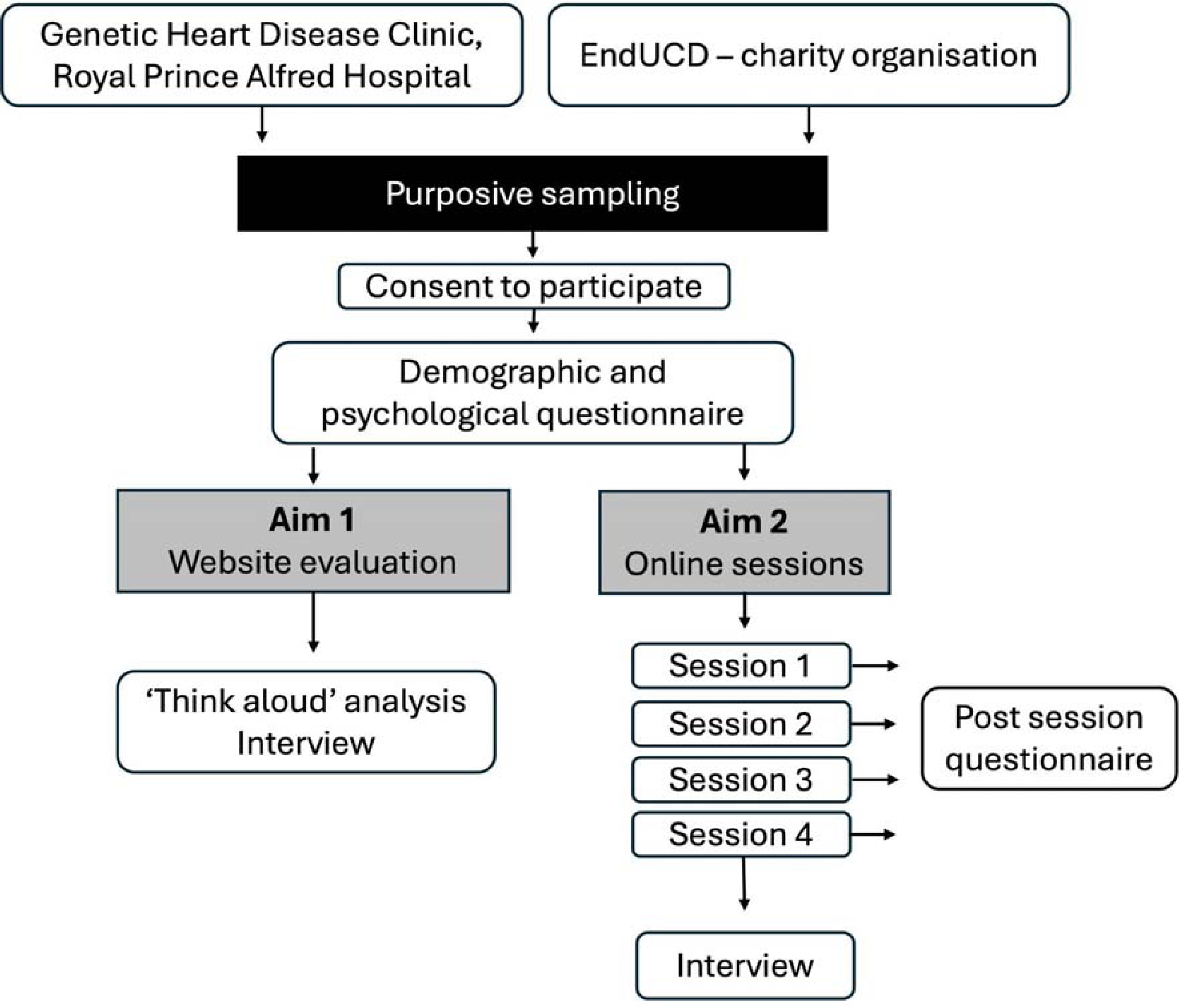
Summary of recruitment and method. UCD = Unexplained cardiac death.

### Baseline survey

Prior to participation, self-reported demographic questions and a series of psychological scales were collected. Demographic questions included ethnicity as classified by one of nine categories from the Australian Bureau of Statistics (*Australian Standard Classification of Cultural and Ethnic Groups (ASCCEG), 2019 | Australian Bureau of Statistics*, 2019).The psychological scales included the Depression Anxiety Stress Scale short form (DASS-21), a validated 21-item scale measuring three subscales depression, anxiety and stress that can be classified into one of five classifications: normal, mild, moderate, severe, extremely severe (Lovibond & Lovibond, 1995); the brief COPE scale (Carver, 1997), a 28-item scale that assesses general life coping over 14 domains, adapted to remove two humour items which was considered insensitive for use among a bereaved population (Buckley et al., 2015); and the Impact of Events Scale-Revised (IES-R), measuring symptoms of post-traumatic stress across three domains (intrusion, avoidance, and hyperarousal) (Weiss & Marmar, 1997). All surveys were completed in REDCap hosted by Sydney Local Health District (Harris et al., 2009, 2019).

### Website evaluation – ‘think aloud’ analysis

Participants underwent a ‘think aloud’ analysis to review the developed website. This qualitative research method asks participants to review content and ‘think aloud’ i.e., verbalise their thoughts while reviewing (Bonner et al., 2014). The ‘think aloud’ analysis was held over video conference using the share screen function and video and audio were recorded. After the participant had reviewed the website, they underwent an interview to further understand their thoughts regarding the website. To assess the acceptability of the website, the interview schedule was designed to address the seven constructs of the theoretical framework of acceptability (Sekhon et al., 2017). A copy of the interview schedule is available in the supplementary information. Transcripts of the interviews were made from the audio recordings but were not made available to the participants for review due to ample opportunity to refine their thoughts throughout the interview. Video recordings were also reviewed to ensure feedback was attributed to the correct section of the website. LY completed all ‘think aloud’ interviews and subsequent analyses.

After completing three ‘think aloud’ interviews, de-identified feedback on each specific page of the website was collated. These were reviewed by two genetic counsellors (LY and AB), and adjustments to the website were made. This included technical issues such as links not working or suggestions to layout/ presentation of the information to improve clarity. Where any discrepancy arose in whether to incorporate changes, these were discussed with the wider research team. An additional three ‘think aloud’ interviews were then completed and proposed adjustments collated and made following the same process.

Transcripts of all six interviews were reviewed to assess the website acceptability. Acceptability was assessed using a deductive framework analysis whereby interview transcripts were reviewed and responses mapped to the construct from the theoretical framework of acceptability. This ensured consideration of all seven constructs. Similar approaches using this framework have been previously reported (Pavlova et al., 2020).

### Online session evaluation

At the completion of each online session, participants completed a short survey that included rating each session out of ten, rating the session on a 5-point Likert scale ranging from very unhelpful to very helpful and giving feedback via open ended questions on elements they liked, and what they found unhelpful.

Participants were invited to undertake a semi-structured interview to provide more feedback. All interviews were conducted by LY over video conference. Interview schedule questions were mapped to the Theoretical Framework of Acceptability (Sekhon et al., 2017). A copy of the interview schedule can be found in the Supplementary Information. All interviews were recorded and transcribed verbatim. A deductive framework analysis was conducted, whereby after familiarisation with the transcripts, responses were mapped to the seven constructs of the theoretical framework of acceptability (Pavlova et al., 2020).

## RESULTS

### Participants

Six of the eight invited participants agreed to participate in the ‘think aloud’ analysis and completed the demographic and baseline questionnaires. Two rounds of ‘think aloud’ analysis were conducted, each with three participants. Twenty clinic patients were invited to participate in the online sessions. Three did not respond and seven declined to participate. Ten individuals from the clinic consented to participate. An additional three participants were recruited through EndUCD. In total, 13 people consented to participate in one of the two groups of four sessions, six in the first group of sessions and seven in the second group of sessions. One participant from group one withdrew after the first session, as they were ‘too overwhelmed’, and were connected to a clinical psychologist. Twelve participants went on to complete the program. The demographics of the participants are shown in Table 1, four individuals participated in both the website review and the online sessions. Two participated in the website review only and the remainder participated in the online sessions only. All participants completed the demographic questionnaire and 14 participants completed the baseline psychological questionnaire. All participants had a DASS-21 depression and anxiety score in the moderate range or above. All participants had a DASS-21 stress score in the mild range or above. In addition, all participants scored an impact of events score above 24 indicating symptoms of post traumatic stress (Table 1). The three highest coping styles as reported by the Brief-COPE scale were: acceptance, active coping and use of emotional support.

**Table 1.** Demographics of the cohort. DASS-21 = Depression, anxiety, stress scale short form.

|  |  |
| --- | --- |
| **n = 15** | **n (%)** |
| Female | 9 (60) |
| University educated | 14 (93) |
| **Study participation** |  |
| Aim 1 – website review only | 2 (13) |
| Aim 2 – online sessions only | 9 (60) |
| Aim 1 and 2 | 4 (27) |
| **Religious affiliation** |  |
| No religion | 8 (53) |
| Christian/ Catholic | 6 (40) |
| Sikh | 1 (7) |
| **Marital status** |  |
| Married | 11 (73) |
| Widowed | 4 (27) |
| **Country of birth** |  |
| Australia | 10 (67) |
| England | 2 (13) |
| Canada | 2 (13) |
| India | 1 (7) |
| **Other medical conditions** | 7 (46)* |
| **Psychological scales (n =14)** |  |
| **Impact of events** | **median (range)** |
| Total | 38 (24-87)** |
| **Impact of events sub scales** | **median (range)** |
| Intrusion | 18 (9-36) |
| Avoidance | 13 (8-29) |
| Hyperarousal | 7 (6-22) |
| **DASS-21 (moderate, severe, extremely severe)** | **n (%)** |
| Depression | 14 (100) |
| Anxiety | 14 (100) |
| Stress | 11 (79) |

### Website content development and readability

A website menu list of content was developed, which brought together similar topics under submenus for ease of navigation (Table 2). Written drafted content was assessed for readability using the Flesch-Kincaid grade level, aiming for a Grade 8 reading level. The mean overall score of the website was 8.75. Some pages were not assessed, namely those that contained (1) clinic contact details, which are unable to be changed, and (2) the family stories page which will be regularly updated and readability assessed with each new story. Where pages scored higher than a Grade 8 level, content was reviewed and simplified.

**Table 2.** Menu of the COPE-SCD website (<u>(Raharjo et al., 2020)</u>) with Flesch-Kincaid grade level of each subpage. NA = not assessed.

| Menu | Submenu | Flesch-Kincaid grade level |
| --- | --- | --- |
| Home page | General information & 'about us' | 9.4 |
| Sudden cardiac death | What is sudden cardiac death | 9.7 |
|  | After the death (coronial process) | 8.7 |
|  | Genetic causes of sudden cardiac death | 9.4 |
|  | Genetic testing | 9.9 |
| Grief and loss | Grief and loss | 9.5 |
|  | Coping strategies | 6.7 |
|  | Support for grieving family and friends | 8.1 |
|  | How to get help | 8.9 |
|  | Unanswered questions | 7.1 |
| Family stories | A place for families to tell their story | NA |
| Group Support | Information on the online support sessions | 8.9 |
| Contact us | Contact information | NA |
|  | List of Genetic Heart Disease Clinics in Australia and New Zealand | NA |

Pages with persistent high scores (e.g., Genetic testing, Flesch-Kincaid score = 9.9), had complicated but essential terms that increased the score e.g., ‘forensic pathologist’, ‘postmortem’ and ‘genetic’. On these pages, a stylistic approach and vector pictures were used to break up the text and enhance understanding. Interactive tools were also used to break text up and focus the reader on the most pertinent information. The website can be viewed at www.copescd.org.au

### ‘Think aloud’ website analysis

Overall, participants liked the website, commenting about the value of the information and the range of topics it covered.

> *“I like the sort of equal emphasis given to both the aspects of sudden cardiac death as well as the grief and loss. Part because they’re both really important and equally pressing factors you have in your mind in a situation like that”*
>
> Participant 5

Participants gave general feedback about the aesthetics of the website, including colour schemes, font sizes and use of stock images. After feedback from the first round of ‘think aloud’ interviews, stock images were removed from the website as they were polarising for the participants with strong opinions in both directions regarding their appropriateness.

These were replaced with simpler vector images, which were well received in the second round. Other concerns participants raised included technical issues, problems with the menu, sections of the website that were missed or overlooked (e.g., video content). Participants also suggested additional content e.g., resources for speaking to children.

### Online session content

Content for the four online sessions is outlined in Table 3. Consideration was given to factors such as allowing time for participants to share their story and for the team to provide information or pictures/ figures to prompt discussion. A copy of the final slide deck for each session can be found in the Supplementary Information.

**Table 3.** Overview of online sessions content.

| Session | Content summary |
| --- | --- |
| One | *Introductory session – introduce participants to the team and each other. Topics covered include:*<br>□ General housekeeping: zoom, respect for other participants, sharing what participants feel comfortable<br>□ Confidentiality<br>□ What to do if feeling unsettled/ need a break<br>□ General information on causes of sudden cardiac death<br>□ Group sharing activity: an opportunity for participants to share their story and ask any questions they may have |
| Two | *Coping with uncertainty – explore the issues of uncertainty after sudden cardiac death particularly when no cause of death is identified. Topics covered include:*<br>□ Coronial process after sudden cardiac death<br>□ When a cause of death is identified vs not identified<br>□ Family screening and genetic testing<br>□ Your family: participants consider their family and first-degree relatives, what tests have they had, do they need additional testing?<br>□ Question and answer session |
| Three | *Coping with grief and loss – with an aim to discuss how grief can manifest and how to cope*<br>□ Discussion of what grief looks like for different people<br>□ Common responses to grief<br>□ Changes in behaviour: withdrawing from people places, avoidance, loss of interest in things.<br>□ Opportunity to share coping strategies and what participants have found helpful or unhelpful |
| Four | *Grief and loss in the wider family – with an aim to recognise how the grief response differs in a family and how it can impact the family unit*<br>□ Normalising differences, reactions and coping among family members<br>□ Positive and negative experiences from family/friends, what's helpful/unhelpful<br>□ Approaching anniversaries, birthdays and missed milestones |

In response to the feedback from participants, iterative changes were made to the content after the completion of the first round of four sessions. Suggested changes included providing more general information on the causes of SCD in session one (moved from session two). In session four, a more inclusive definition of what is meant by “family”, was suggested recognising that when considering grief responses, “family” is not limited to your genetic first-degree relatives, but also needs to consider those around you in the community who provide support. No further suggestions for change in content were made in the second set of sessions, however there was feedback about the length of the sessions, this will be discussed further below.

### Online session feedback

Post-session questionnaire responses were collated for each session, overall participants responded positively to each session. Supplementary Table S1 summarises the scores for each session and gives an example quote for the open-ended questions.

Nine participants elected to undertake the post-session interview, four from the first group of sessions and five from the second group. Peer support was a benefit noted by most participants:

> *“I think they [the online sessions] just validated how I was feeling and what I’d been feeling … that yeah. Wasn’t, wasn’t going crazy. <laugh>, I’m not the only one”*.
>
> Participant 6

> *“There’s, uh, a lot of strength to be found in numbers [peer support], if you like, and, and people saying the same thing”.* Participant 11

Participants valued hearing the perspectives of a variety of people. Particularly hearing the opinions of those who had a different relationship to the deceased helped them as they considered the responses of different family members, yet they noted that this required sensitivity from the participants.

> *“I liked the fact that you had parents, siblings and spouses involved. Because I thought that provided a variety of different perspectives. Um, and it particularly helps me as a parent in terms of my son about how he might feel about different things. So, understanding his perspective a little bit more.”* Participant 12.

> *“Sometimes it felt like there was that disconnect between people who had lost a spouse as opposed to people who had lost a child as opposed to people who had lost a sibling, um, and not understand, not fully understanding what they’re going through. So, in a way it, I think it was good to understand where they’re coming from and hearing what they were saying, but also, yeah, I think just again, that sensitivity that other people grieve in different ways”* Participant 8

Efforts were made to evaluate the ideal group size, with more participants invited after the first round. However, participants from the second group of sessions noted time was a limiting factor when considering the number of participants, and it was important to ensure ample time for each participant to contribute. In the second group of sessions, participant numbers were increased to seven, however interview feedback highlighted that for sessions three and four, because of their focus on grief and loss and sharing around coping, an hour was insufficient time.

> *“I think the sessions probably could have gone for longer. So, I think an hour and a half would feel right. I think if it’s at two hours it’s probably a bit too much of a time commitment for people to agree to … an hour and a half probably would work”* Participant 13

### Acceptability assessment

Participants found both the website and the online sessions to be acceptable as demonstrated by mapping their responses to the theoretical framework of acceptability (Sekhon et al., 2017). Example quotes from the evaluation of the website and the online sessions are outlined in Table 4.

**Table 4.** Example quotes from the website review and online sessions addressing the acceptability constructs of the COPE-SCD intervention. P = participant.

| Acceptability construct | Construct description | Website review | Online session |
| --- | --- | --- | --- |
| Affective attitude | How an individual feels about the intervention | *"having sort of one place to go for all the information is really good"* P14 | *"I just found...sharing and listening to the other people and their views and how they're coping with their grief...I found that really valuable"* P6 |
| Burden | The perceived amount of effort that his required to participate | *"I think that most of the information is presented in a fairly easily accessible way"* P15 | *"And it's very convenient [zoom] I suppose when you've got people that are spread far and wide. I gather it's probably easier to do it that way than trying to do it in person"* P11 |
| Ethicality | The extent to which the intervention has a good fit with an individual's value system | *"There was nothing there that kind of, I found offensive or, unduly upsetting"* P1 | *"I think it like aligned with my values fairly well and then I think everyone was really respectful towards each other"* P4 |
| Opportunity costs | The extent to which benefits, profits or values must be given up to engage in the intervention | *"It would be a very good use of my time"* P15 | *"I would encourage anybody that finds themselves in this situation to do something, a workshop like that...I've gained a lot more out of this kind of a workshop as opposed to one-to-one counselling"* P11 |
| Intervention coherence | The extent to which the participant understands the intervention and how it works | *"Something like this early on, it'd be helpful for me, um, to answer some of those questions, those common questions, but then also to share it say on social media, on Facebook and say, you know, I have found this really helpful"* P6 | *"it was easy to get into that routine so I knew, you know, four o'clock every Wednesday I've got this session, it goes for an hour"* P8 |
| Perceived effectiveness | The extent to which the intervention is perceived as likely to achieve its purpose | *"I think it, it catches a few of those really big feelings and questions that you have in this situation"* P5 | *"I guess it allowed me to probably look back a little bit in time in terms of what I've been through...wow, [participant], that's really good. You've come through that you've managed okay"* P12 |
| Self-efficacy | The participants confidence that they can perform the behaviour(s) required | *"it's ease of navigation. It was simple"* P4 | *"I think just overall, it, it just reassured me that my own reactions, my own way of, uh, my own way of dealing with it on a day-to-day basis are very similar to everybody else's"* P11 |

## DISCUSSION

Supporting families after SCD is a key aspect of cardiovascular genetic counseling practice and evidence-based tools and approaches to do this are critically needed. Families experience marked psychological distress (Bates et al., 2019; Ingles et al., 2016; Yeates et al., 2013) and have expressed a need for more information and support (McDonald et al., 2020; Steffen et al., 2020). Numerous guidelines highlight the need for psychological support for these families and most services have insufficient support options available (*2023 ESC Guidelines for the Management of Cardiomyopathies | European Heart Journal | Oxford Academic*, n.d.; Stiles et al., 2021; van den Heuvel et al., 2024). In Australia, barriers exist to accessing mental healthcare providers and these are exacerbated in rural communities (Kavanagh et al., 2023). In addition, SCD families request support from professionals with expertise in the nuances of SCD (van den Heuvel et al., 2024). A scalable solution is needed and in response, we developed COPE-SCD: an online community supporting families after SCD in the young, consisting of a website and online sessions(Yeates et al., 2022). COPE-SCD is the first Australian intervention to provide accessible information and support specifically to families who have suffered a young SCD. The intervention was found to be acceptable when assessed using the Theoretical Framework of Acceptability (Sekhon et al., 2017) and will undergo subsequent evaluations to determine effectiveness in improving psychological outcomes.

Implementation research suggests review of acceptability both in the early stages of intervention development as well as once it is integrated into use (Klaic et al., 2022). Perski and Short argue that acceptability should also be considered as part of a broader system that associates closely with user engagement and intervention effectiveness (Perski & Short, 2021). Participants reported the convenience of the online session that allows real time interactions with facilitators and co-participants (assessed under the burden and intervention coherence constructs). One potential negative of an internet-based peer support model is the removal of “incidental” discussions before and after the formal session, and this was raised by our participants. Indeed, some mentioned the time at the completion of the session where one might stay back and continue to discuss topics with other participants was not possible, and they were left sitting alone in a room after having engaged in emotive discussions. More work is needed to explore ways to recreate personal engagement and incidental conversations in the online environment.

A key outcome of previous work (van den Heuvel et al., 2024) and the codesign stakeholder feedback that underpinned this intervention (Yeates et al., 2022), was the lack of information available to families on causes of SCD, next steps for family members, and grief and loss.

The website component is a key provider of information in the COPE-SCD intervention. However, we also recognise the benefit and opportunity the online sessions provided for information, discussion and questions. This format may also be better suited to those with low written literacy levels and language barriers where spoken information complimented by visual aids is of more benefit. The content covered and discussion topics were designed to be moderated by healthcare professionals, though it is important to consider whether the sessions, particularly sessions three and four, could be moderated by a trained “peer”. This would reduce time burden on the healthcare team and may create a more comfortable space for participants. This type of peer-led support group has been used in bereavement after SCD (Steffen et al., 2020).

In the grief and bereavement setting, peer support has shown benefit in different clinical scenarios, including death due to suicide (Oulanova et al., 2014) and long-term illness such as cancer (Raharjo et al., 2020). Other studies in the setting of SCD have reported a preference for peer support (Steffen et al., 2020; van den Heuvel et al., 2024) and our participants valued the opportunity to share similar experiences (Yeates et al., 2022). A systematic review found that of the studies reviewed, over half reported benefits for peer support including a reduction of grief symptoms such as depression and isolation (Bartone et al., 2019).

Meaning making, which is an important concept in grief and loss, is an intrinsic part of human nature that resolves to understand and anticipate events in our life and find meaning in them (Neimeyer, 2019). Following the SCD of a young relative, the bereaved embark on new meaning making which can be particularly problematic given there is often so much uncertainty (Neimeyer et al., 2014). “Making sense” of why the young person passed away or the “desire to know” the cause of death is commonly expressed by SCD family members (Steffen et al., 2020; van den Heuvel et al., 2024). Online session two covers the causes of SCD in young people and addresses managing uncertainty when no cause of death has been identified. Neimeyer et al., (2014) argue that narration, as exhibited in storytelling, is a key part of meaning making in bereavement (Neimeyer et al., 2014). Session one provides an opportunity for participants to tell their story and promotes an opportunity for narration.

### Limitations

The majority of participants were recruited through a tertiary referral centre with extensive experience in caring for families after SCD and likely represent a more well-adjusted sub-group. Our participants were mainly university educated and of European background, lacking diversity. When undertaking effectiveness studies, future efforts to recruit participants from more diverse backgrounds and clinical experiences is needed to ensure the program has broad applicability. The ‘think aloud’ analysis was based on interviews with six participants, this number is consistent with other studies using ‘think aloud’ methods to assess health interventions (Bonner et al., 2014; Jaspers et al., 2004). However, we recognise the subjective nature of a website, where 100% agreement is unlikely to be reached, regardless of the sample size.

### Practice implications

The COPE-SCD intervention is an Australian-first for providing online information and psychological support to families after SCD in the young. We have shown this codesigned intervention to be acceptable and future work will evaluate effectiveness with an aim to implement this intervention into clinical care. The program provides a new resource for genetic counselors supporting families after SCD. The versatility of an online program is that it could be easily adjusted to the local context of other clinical services located nationally and internationally, recognising that regional differences do exist.

## CONCLUSION

We describe the development and acceptability assessment of COPE-SCD: a support intervention for families affected by SCD in the young. The intervention includes a website, which provides information in written and video format on both causes of SCD in young people and coping with grief and loss. The second part of the intervention, the online sessions, builds on the information provided on the website and provide an opportunity for peer support. We assessed acceptability of the intervention using Sekhon’s Theoretical Framework of Acceptability. Overall, participants found the intervention acceptable when considering seven constructs of acceptability. More work is needed to assess the effectiveness of the intervention as it’s implemented into clinical practice. It is our hope the COPE-SCD will provide much needed support for families after SCD in the young.

## Data Availability

Anonymized data can be made available upon reasonable request and with appropriate agreements and human research ethics committee approval.

## REFERENCES

2023 ESC Guidelines for the management of cardiomyopathies | European Heart Journal | Oxford Academic. (n.d.). Retrieved March 24, 2025, from https://academic.oup.com/eurheartj/article/44/37/3503/7246608

Australian Standard Classification of Cultural and Ethnic Groups (ASCCEG), 2019 | Australian Bureau of Statistics. (2019, December 18). https://www.abs.gov.au/statistics/classifications/australian-standard-classification-cultural-and-ethnic-groups-ascceg/latest-release

Ayre, J., Bonner, C., Muscat, D. M., Dunn, A. G., Harrison, E., Dalmazzo, J., Mouwad, D., Aslani, P., Shepherd, H. L., & McCaffery, K. J. (2023). Multiple Automated Health Literacy Assessments of Written Health Information: Development of the SHeLL (Sydney Health Literacy Lab) Health Literacy Editor v1. JMIR Formative Research, 7(1), e40645. 10.2196/40645

Bagnall, R. D., Weintraub, R. G., Ingles, J., Duflou, J., Yeates, L., Lam, L., Davis, A. M., Thompson, T., Connell, V., Wallace, J., Naylor, C., Crawford, J., Love, D. R., Hallam, L., White, J., Lawrence, C., Lynch, M., Morgan, N., James, P.,… Semsarian, C. (2016). A Prospective Study of Sudden Cardiac Death among Children and Young Adults. New England Journal of Medicine, 374(25), Article 25. 10.1056/NEJMoa1510687

Bartone, P. T., Bartone, J. V., Violanti, J. M., & Gileno, Z. M. (2019). Peer Support Services for Bereaved Survivors: A Systematic Review. OMEGA - Journal of Death and Dying, 80(1), Article 1. 10.1177/0030222817728204

Bates, K., Sweeting, J., Yeates, L., McDonald, K., Semsarian, C., & Ingles, J. (2019). Psychological adaptation to molecular autopsy findings following sudden cardiac death in the young. Genetics in Medicine, 21(6), Article 6. 10.1038/s41436-018-0338-4

Bonner, C., Jansen, J., Newell, B. R., Irwig, L., Glasziou, P., Doust, J., Dhillon, H., & McCaffery, K. (2014). I Don’t Believe It, But I’d Better Do Something About It: Patient Experiences of Online Heart Age Risk Calculators. Journal of Medical Internet Research, 16(5), e3190. 10.2196/jmir.3190

Buckley, T., Spinaze, M., Bartrop, R., McKinley, S., Whitfield, V., Havyatt, J., Roche, D., Fethney, J., & Tofler, G. (2015). The nature of death, coping response and intensity of bereavement following death in the critical care environment. Australian Critical Care, 28(2), Article 2. 10.1016/j.aucc.2015.02.003

Caleshu, C., Kasparian, N. A., Edwards, K. S., Yeates, L., Semsarian, C., Perez, M., Ashley, E., Turner, C. J., Knowles, J. W., & Ingles, J. (2016). Interdisciplinary psychosocial care for families with inherited cardiovascular diseases. Trends in Cardiovascular Medicine, 26(7), Article 7. 10.1016/j.tcm.2016.04.010

Cardiac Risk in the Young. (2024, June 10). https://www.c-r-y.org.uk/

Carver, C. S. (1997). You want to measure coping but your protocol’s too long: Consider the brief COPE. International Journal of Behavioral Medicine, 4(1), Article 1. 10.1207/s15327558ijbm0401_6

Cheng, C., & Dunn, M. (2015). Health literacy and the Internet: A study on the readability of Australian online health information. Australian and New Zealand Journal of Public Health, 39(4), 309–314. 10.1111/1753-6405.12341

Coffey, T., Duncan, E., Morgan, H., & Gillies, K. (2023). Developing strategies to address disparities in retention communication during the consent discussion: Development of a behavioural intervention. Trials, 24(1), Article 1. 10.1186/s13063-023-07268-2

Harris, P. A., Taylor, R., Minor, B. L., Elliott, V., Fernandez, M., O’Neal, L., McLeod, L., Delacqua, G., Delacqua, F., Kirby, J., & Duda, S. N. (2019). The REDCap consortium: Building an international community of software platform partners. Journal of Biomedical Informatics, 95, 103208. 10.1016/j.jbi.2019.103208

Harris, P. A., Taylor, R., Thielke, R., Payne, J., Gonzalez, N., & Conde, J. G. (2009). Research electronic data capture (REDCap)—A metadata-driven methodology and workflow process for providing translational research informatics support. Journal of Biomedical Informatics, 42(2), 377–381. 10.1016/j.jbi.2008.08.010

Ingles, J., & James, C. (2017). Psychosocial care and cardiac genetic counseling following sudden cardiac death in the young. Progress in Pediatric Cardiology, 45, 31–36. 10.1016/j.ppedcard.2017.03.001

Ingles, J., Spinks, C., Yeates, L., McGeechan, K., Kasparian, N., & Semsarian, C. (2016). Posttraumatic Stress and Prolonged Grief After the Sudden Cardiac Death of a Young Relative. JAMA Internal Medicine, 176(3), Article 3. 10.1001/jamainternmed.2015.7808

Jaspers, M. W. M., Steen, T., Bos, C. van den, & Geenen, M. (2004). The think aloud method: A guide to user interface design. International Journal of Medical Informatics, 73(11), Article 11. 10.1016/j.ijmedinf.2004.08.003

Kavanagh, B. E., Corney, K. B., Beks, H., Williams, L. J., Quirk, S. E., & Versace, V. L. (2023). A scoping review of the barriers and facilitators to accessing and utilising mental health services across regional, rural, and remote Australia. BMC Health Services Research, 23(1), 1060. 10.1186/s12913-023-10034-4

Klaic, M., Kapp, S., Hudson, P., Chapman, W., Denehy, L., Story, D., & Francis, J. J. (2022). Implementability of healthcare interventions: An overview of reviews and development of a conceptual framework. Implementation Science, 17(1), 10. 10.1186/s13012-021-01171-7

Lovibond, S., & Lovibond, P. (1995). Manual for Depression anxiety stress scale (Second edition).

Marijon, E., Narayanan, K., Smith, K., Barra, S., Basso, C., Blom, M. T., Crotti, L., D’Avila, A., Deo, R., Dumas, F., Dzudie, A., Farrugia, A., Greeley, K., Hindricks, G., Hua, W., Ingles, J., Iwami, T., Junttila, J., Koster, R. W.,… Winkel, B. G. (2023). The *Lancet* Commission to reduce the global burden of sudden cardiac death: A call for multidisciplinary action. The Lancet, 402(10405), 883–936. 10.1016/S0140-6736(23)00875-9

McDonald, K., Sharpe, L., Yeates, L., Semsarian, C., & Ingles, J. (2020). Needs analysis of parents following sudden cardiac death in the young. Open Heart, 7(2), Article 2. 10.1136/openhrt-2019-001120

Neimeyer, R. A. (2019). Meaning reconstruction in bereavement: Development of a research program. Death Studies, 43(2), Article 2. 10.1080/07481187.2018.1456620

Neimeyer, R. A., Klass, Dennis, & and Dennis, M. R. (2014). A Social Constructionist Account of Grief: Loss and the Narration of Meaning. Death Studies, 38(8), 485–498. 10.1080/07481187.2014.913454

Oulanova, O., Moodley, R., & Séguin, M. (2014). From Suicide Survivor to Peer Counselor: Breaking the Silence of Suicide Bereavement. OMEGA - Journal of Death and Dying, 69(2), 151–168. 10.2190/OM.69.2.d

Pavlova, N., Teychenne, M., & Olander, E. K. (2020). The Concurrent Acceptability of a Postnatal Walking Group: A Qualitative Study Using the Theoretical Framework of Acceptability. International Journal of Environmental Research and Public Health, 17(14), Article 14. 10.3390/ijerph17145027

Paynter, C., McDonald, C., Story, D., & Francis, J. J. (2023). Application of the theoretical framework of acceptability in a surgical setting: Theoretical and methodological insights. British Journal of Health Psychology, 28(4), 1153–1168. 10.1111/bjhp.12677

Perski, O., & Short, C. E. (2021). Acceptability of digital health interventions: Embracing the complexity. Translational Behavioral Medicine, 11(7), Article 7. 10.1093/tbm/ibab048

Raharjo, C. V., Hetherington, K., Donovan, L., Fardell, J. E., Russell, V., Cohn, R. J., Morgan, N. L., Siddiqui, J., & Wakefield, C. E. (2020). An Evaluation of By My Side: Peer Support in Written Form is Acceptable and Useful for Parents Bereaved by Childhood Cancer. Journal of Pain and Symptom Management, 59(6), 1278–1286. 10.1016/j.jpainsymman.2020.01.013

Sekhon, M., Cartwright, M., & Francis, J. J. (2017). Acceptability of healthcare interventions: An overview of reviews and development of a theoretical framework. BMC Health Services Research, 17(1), Article 1. 10.1186/s12913-017-2031-8

Steffen, E. M., Timotijevic, L., & Coyle, A. (2020). A qualitative analysis of psychosocial needs and support impacts in families affected by young sudden cardiac death: The role of community and peer support. European Journal of Cardiovascular Nursing, 19(8), Article 8. 10.1177/1474515120922347

Stiles, M. K., Wilde, A. A. M., Abrams, D. J., Ackerman, M. J., Albert, C. M., Behr, E. R., Chugh, S. S., Cornel, M. C., Gardner, K., Ingles, J., James, C. A., Juang, J.-M. J., Kääb, S., Kaufman, E. S., Krahn, A. D., Lubitz, S. A., MacLeod, H., Morillo, C. A., Nademanee, K.,… Wang, D. W. (2021). 2020 APHRS/HRS expert consensus statement on the investigation of decedents with sudden unexplained death and patients with sudden cardiac arrest, and of their families. Journal of Arrhythmia, 37(3), Article 3. 10.1002/joa3.12449

Sudden Arrhythmia death syndromes (SADS) foundation. (n.d.). https://sads.org/

Turnbull, S., Walsh, N. E., & Moore, A. J. (2023). Adaptation and Implementation of a Shared Decision-Making Tool From One Health Context to Another: Partnership Approach Using Mixed Methods. Journal of Medical Internet Research, 25, e42551. 10.2196/42551

van den Heuvel, L., Do, J., Yeates, L., Burns, C., Semsarian, C., & Ingles, J. (2024). Sudden cardiac death in the young: A qualitative study of experiences of family members with cardiogenetic evaluation. Journal of Genetic Counseling, 33(2), 361–369. 10.1002/jgc4.1733

van der Houwen, K., Stroebe, M., Schut, H., Stroebe, W., & van den Bout, J. (2010). Online mutual support in bereavement: An empirical examination. Computers in Human Behavior, 26(6), Article 6. 10.1016/j.chb.2010.05.019

Weiss, D., & Marmar, C. (1997). The impact of event scale—Revised. In Assessing psychological trauma and PTSD. (pp. 399–411). Guilford Press.

Yeates, L., Gardner, K., Do, J., van den Heuvel, L., Fleming, G., Semsarian, C., McEwen, A., Adlard, L., & Ingles, J. (2022). Using codesign focus groups to develop an online COmmunity suPporting familiEs after Sudden Cardiac Death (COPE-SCD) in the young. BMJ Open, 12(8), Article 8. 10.1136/bmjopen-2021-053785

Yeates, L., Hunt, L., Saleh, M., Semsarian, C., & Ingles, J. (2013). Poor psychological wellbeing particularly in mothers following sudden cardiac death in the young. European Journal of Cardiovascular Nursing, 12(5), Article 5. 10.1177/1474515113485510

