## Supplementary information for "Development and acceptability of a support intervention for families after sudden cardiac death in the young"

Laura Yeates BSc (hons) Grad Dip Gen Couns PhD [1-3], Amy Baker BSc, MGenCouns[1], Karen Gardner BA, MPH, PhD, Natalie Stewart MGenCouns[1], Laura Catto MGenCouns [1], Judy Do BPsych (Hons)[1], Sue Timbs BCom LLB6, Felicity Leslie BAppSc (MRT) [1], Christopher Semsarian MBBS, PhD, MPH[2,3,7], Belinda Gray MBBS PhD[2,3], Leesa Adlard MA ClinPsych, D.Litt et Phil Psych[3], Jodie Ingles GradDipGenCouns, PhD, MPH[1-3]

[1] Genomics and Inherited Disease Program, Garvan Institute of Medical Research, and UNSW Sydney, Sydney, Australia

[2]Faculty of Medicine and Health, The University of Sydney, Sydney, Australia

[3]Department of Cardiology, Royal Prince Alfred Hospital, Sydney, Australia

[4]Academic Unit of General Practice, Australian National University, Canberra, Australia

[6]End unexplained cardiac death (EndUCD.org)

[7]Agnes Ginges Centre for Molecular Cardiology at Centenary Institute, The University of Sydney, Sydney, Australia

#### Supplementary information

**COPE-SCD "Think Aloud" interview schedule**

Thank you for agreeing to be interviewed. As you know we are performing a study to evaluate a new intervention to support families after a sudden cardiac death in the young.

This part of the study focuses specifically on evaluating a new website. Thank you for completing the online consent form and baseline survey.

**Confidentiality/recording:**
Everything you say in this interview will be strictly confidential. Your name and any identifying details will be removed from the written transcript, which will be made from an audio recording of the interview.

We understand that reviewing a website focused on sudden cardiac death in young people may be confronting, if you'd prefer not to answer any questions please just let me know and we'll move to the next question. You can also stop the interview at any time.

The first part of this interview is a "think aloud" analysis, this is where we need you to talk out aloud as you review our website, giving your opinion on the website. There's no right or wrong answers, we're keen to get your honest opinions on layout, technical difficulties and content.

In a moment I will share two websites with you, the first is a warm up exercise to get you used to "thinking aloud", once you've had a practice for a few minutes then we'll move to the study website.

As we go through if you're silent for more than 10 seconds I will prompt you to "think aloud".

Once you've finished reviewing the website there'll be a few additional questions at the end. As well as a short usability survey.

Does that sound ok? Do you have any questions?

*****START RECORDER*****

---

If you can please open your web browser and go to www.spotthedifference.com

Now press "share screen". You'll see there are two pictures please think aloud while you "spot the difference"

*[Prompt keep talking as you go]*

Great thank you, now you have the hang of it, I'll now send you the link to the website via the chat function.

If you could please review all parts of the website, including any video content and think aloud as you go. I would suggest reviewing each page from top to bottom, and then the pages along the menu from right to left.
*[prompts: keep thinking aloud, if having difficulty navigating the website provide as little assistance as possible.*

Once they have finished reviewing the website, ask them to “unshare screen” and close the webpage.

**Post website review questions:**

1. Thank you for doing all of that, how did you find that process in general?
2. What did you like the best about the website?
3. What did you like the least about the website?
   How did the website make you feel?
   a. prompts: friendly? clinical?
4. How easy or hard was it to navigate the website?
   Were there any parts you found frustrating?
   Was there any content you found hard to read?

   a. *[Prompt - perhaps it was worded poorly, or the information made you feel uncomfortable?]*

5. Was the language easy to read and understand/ too technical/ emotionally difficult to read
6. Do you think the website aligned with your value system i.e. were you offended by any of its content?
   Do you think this website would be helpful to a family affected by sudden cardiac death?
7. Can you tell me a little bit more about why you answered this way? Can you tell me more about what was helpful/ not helpful?
8. What would you score the website out of 10?
9. Do you think reviewing this website was a good use of your time?
10. Do you think you could navigate around the website on your own, i.e. could you find specific content again?
11. Did you have a preference for video or written content?
    Was there too much text or not enough?
    Do you have any other suggestions for how the website could be improved?
12. Is there anything else you would like to comment on that we haven’t covered?

**Thank you for your time.**

**STOP RECORDING**

**COPE SCD - Post Online session interview schedule**

Thank you for agreeing to be interviewed. As you know we are performing a study to evaluate a new intervention to support families after a sudden cardiac death in the young.

This part of the study focuses specifically on evaluating the series of four online sessions.

**Confidentiality/recording:**
Everything you say in this interview will be strictly confidential. Your name and any identifying details will be removed from the written transcript, which will be made from an audio recording of the interview.

We understand speaking about your experiences and participation in the program may cause some distress, if you'd prefer not to answer any questions please just let me know and we'll move to the next question. You can also stop the interview at any time.

Do you have any questions before we start?

*****START RECORDER*****

---

1. Overall, how did you find the online sessions? Which parts did you like the best, what did you like the least.
2. Overall, how did participating in the online sessions make you feel? Clarifying: did you worry about attending, did you look forward to attending?
3. How did you find the online format over Zoom? Was it easy to use?
4. How did you find the amount of time and effort to participate?
    a. Prompts - about right, too much, too little
5. How did you find the time between sessions?
    a. Prompts - about right, too long, too short
6. How did you feel in the 4 weeks during the sessions and between the sessions?
    a. Were you nervous or looking forward to each session?
    b. Did you find the sessions particularly draining or did they give you energy?
    c. How did you feel you coped between sessions? What was hard, what was easy?
    d. Did you find yourself thinking about the previous session a lot between sessions?
7. Can you think about your values or what is important to you? Values can include your principles, standards, morals, religious views etc.
    a. How did the sessions align/ not align with your values?
    b. Did you find yourself agreeing/disagreeing with the content?
    c. On balance, do you think participating in the online sessions was a good use of your time?

    d. Are there other ways you could have used your time that would have been more beneficial to you that would support you after a sudden death in the family?
8. How did the online sessions help/ not help you understand your experience better?
9. Did the online sessions help you to understand/ better empathise with your family members?
    a. If yes how
    b. if no, do you have any suggestions on why they didn't help?
10. Were there any particular "lessons learnt" or take home messages that have helped you and your family to cope?

11. Do you feel confident to apply some of these ideas discussed to your situation/ family?
    a. Is so which ones,
    b. if not can you tell me a bit more about why that wouldn't work for your family
12. What was the most valuable part of the online course for you?
13. Is there anything else you would like to add?

**Thank you for your time!**

**STOP RECORDER**

**Slide decks from the four COPE-SCD online sessions (notes to help guide the facilitator are under each slide)**

**SESSION 1**

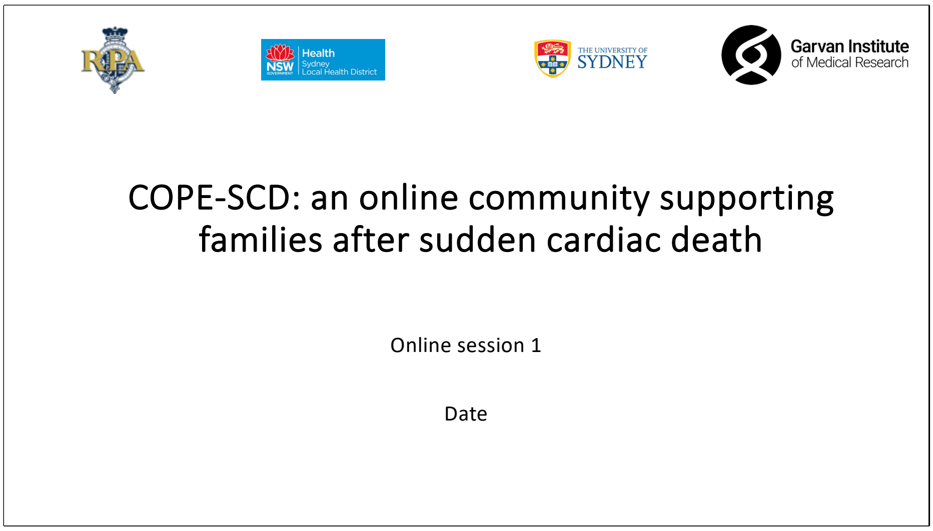

Garvan Institute of Medical Research

Welcome!

#### Meet the team

Garvan Institute of Medical Research

- Laura Yeates (Genetic Counsellor)
- Dr Leesa Adlard (Clinical Psychologist)
- A/Prof Jodie Ingles (Genetic Counsellor)
- Prof Chris Semsarian (Cardiologist)
- Dr Belinda Gray (Cardiologist)

Introduce each member of the team (not all need to be in attendance for every session).

#### Background – Why do this?

Garvan Institute of Medical Research

- In working with families affected by sudden cardiac death, we recognise lots of difficulties
- In response to this we conducted focus groups to develop a support intervention for families after sudden cardiac death

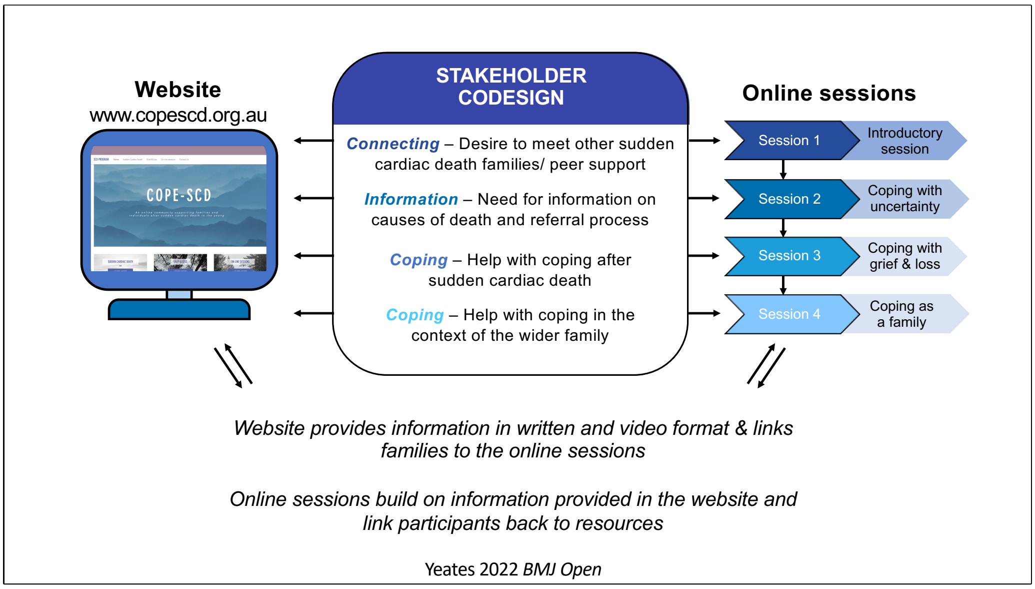


Main themes of the codesign process are shown in centre of this diagram. Families told us their main needs were connecting with other families/ peer support, information on causes of sudden cardiac death and help with coping. Coping on both a personal level and in the wider context as a family.

We've used the findings of this study to structure the sessions.

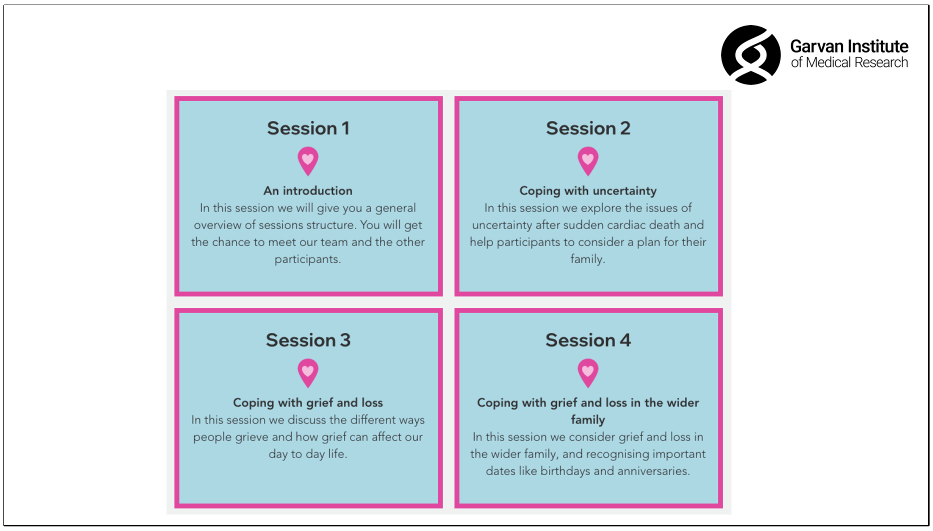


This slide will be repeated each session and just gives context on what today's session will be on and where we'll head next week.

#### Housekeeping

- **Confidentiality** – after the session if you're sharing with a friend about what was discussed, please don't use names
- **Selfcare** – Some of what you hear might trigger emotions, you can turn off your camera, go get some water, stand up etc.
- **Peer support** is a key part of this program – sharing experiences is helpful

### Plan for today

Garvan Institute of Medical Research

- A few facts on sudden cardiac death
- Introductions
  - A chance to get to know each other
- What are you hoping to get out of the program?

### Some statistics

Garvan Institute of Medical Research

- In Australia & New Zealand 2 - 3 young people age 1-35 years die suddenly each week[1]
- 40% of young people who die suddenly have no cause identified at post-mortem[1]
- When no cause of death is identified, genetic testing will identify a cause of death in about 20% of families[1,2]


1. Bagnall et al 2016 *NEJM*
2. Isbister et al 2021 *Int J Cardiology*

#### Some statistics

- In all age groups, one study reported 77% of people who suffer a cardiac arrest have no prior symptoms
- The chance of surviving a cardiac arrest outside of hospital is less than 10%[1]

***More information on this next week***

1. Paratz et al. 2020. *Heart rhythm*

#### Introductions

- Tell us a bit about your family?
- How long ago your family member passed away?
- A little bit about them, e.g. favourite hobby or sport

It's important to give everyone a chance to tell their story. It doesn't have to be long, just what they feel comfortable in sharing.

Recognising there are many reasons to join these sessions can you tell us why you're here ?(poll)

- To meet other families
- For medical information
- To find out about genetic testing
- To help other families
- To help me cope
- To help my family cope
- Something different?

We set this up as a poll in zoom and then shared the responses with the participants.

#### Thank you for attending!

Garvan Institute of Medical Research

- After today's session:
  - What are you going to do to look after yourself now?
  - If you feel distressed, who can you talk to? (family, friend, Beyond Blue – 1300 22 4636)
  - Please contact our team if you need further assistance
  - Any questions or anything you'd like to add?

This slide is repeated at the end of each session as a prompt to consider how the participants are going to care for themselves now recognizing the discussion may have brought up some emotions for them.

#### Dates for your diary

Garvan Institute of Medical Research

- Session 2 – date and time
- Session 3 – date and time
- Session 4 – date and time

### SESSION 2

COPE-SCD: an online community supporting families after sudden cardiac death

Online session 2

Date

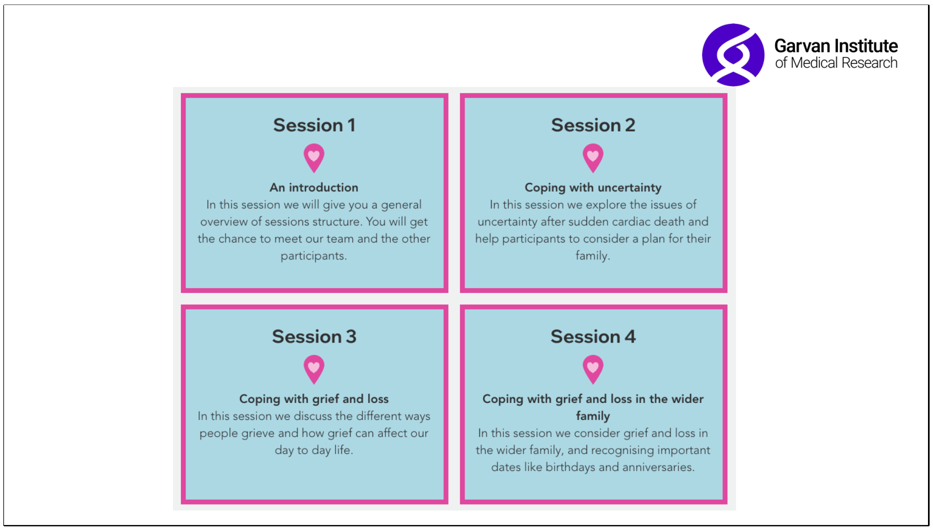


This slide will be repeated each session and just gives context on where today's session fits in and where we'll head next week.

### Housekeeping

Garvan Institute of Medical Research

- **Confidentiality** – after the session if you're sharing with a friend about what was discussed, please don't use names
- **Selfcare** – Some of what you hear might trigger emotions, you can turn off your camera, go get some water, stand up etc.
- **Peer support** is a key part of this program – sharing experiences is helpful

### Checking in after session 1

- How did you go after last week?
- How did you look after yourself after the session?

This is will also be repeated in sessions 2-4, to allow space for people to reflect or ask any questions after the previous week.

### Plan for today

Garvan Institute of Medical Research

- What happens after sudden cardiac death?
- Q and A

Outline of current session.

#### Caring for the family after sudden death

Garvan Institute of Medical Research

**Investigation of the Decedent**
*Circumstances of death, previous illness, family history etc*

**Review of Post-Mortem Findings**
*Was a cause identified? Was there "no cause"?*

**1. Clinical Evaluation of Family**
*Usually start with "first-degree relatives". ECG, Echo, Ex test etc*

**2. Genetic Analysis of Post-Mortem Sample**
*May be helpful in 15-20% of families*

**3. Psychological Support for Family**
*Prolonged grief, anxiety, depression, PTSD etc*

How we care for the family after sudden cardiac death.
Initially (steps 1 and 2) we're determining is there a possible genetic cause of death. If there is a possible genetic cause, then we move to the numbered points in how we look after family members.

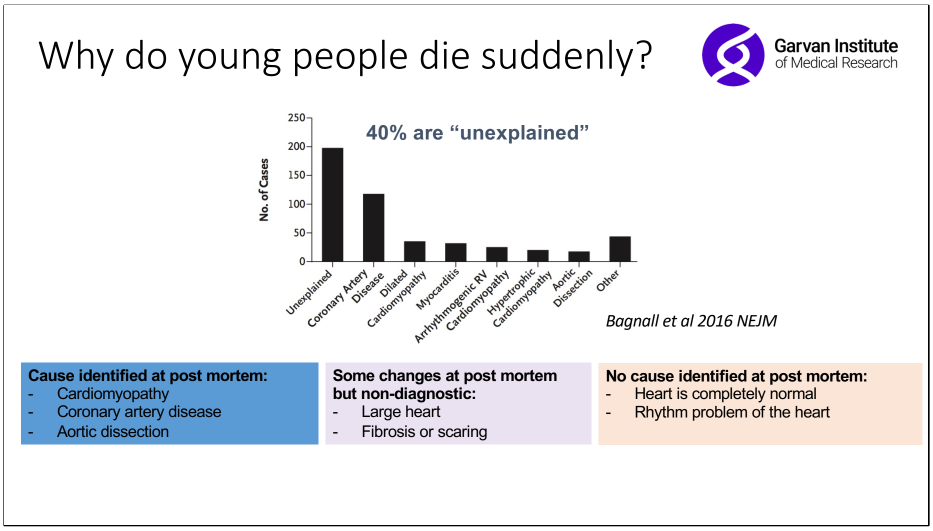


Looking at the causes of sudden cardiac death in the young. Graph from Bagnall et al 2016 NEJM https://doi.org/10.1056/NEJMoa1510687
Broadly speaking three possible categories of what might be identified at post mortem with examples.

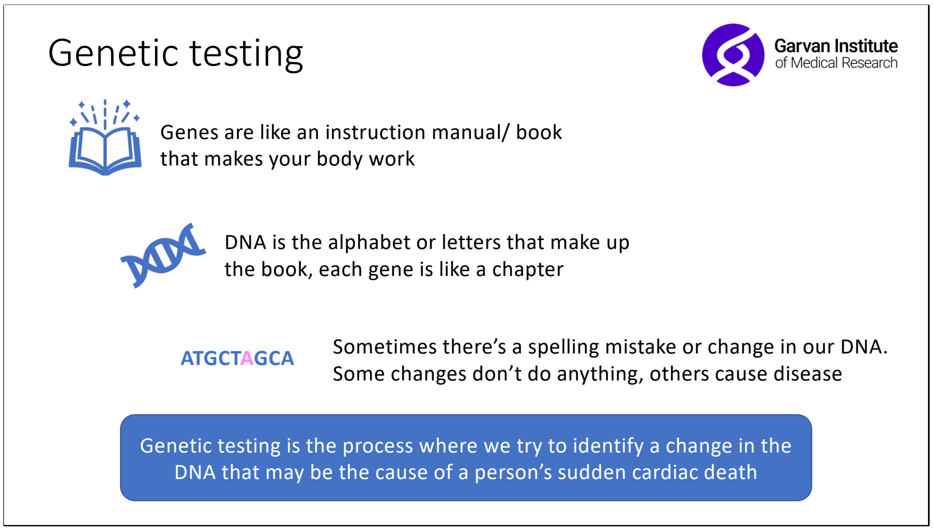


Overview of the process of genetic testing

### Genetic testing

Garvan Institute of Medical Research

- Requires a DNA sample of good quality
- Can take some time depending on the type of testing ordered
- Interpretation of results can be difficult

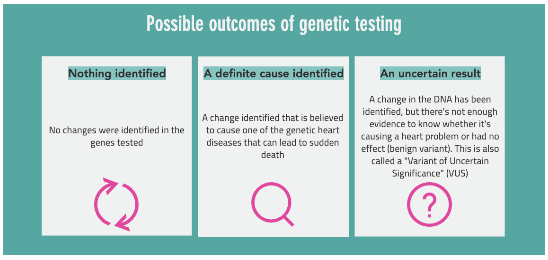


The availability of a post mortem sample for genetic testing will depend on where the death occurred and how long ago they passed away. In Australia, most young people who die suddenly have a blood sample stored. This has been mandated as best practice since 2008.

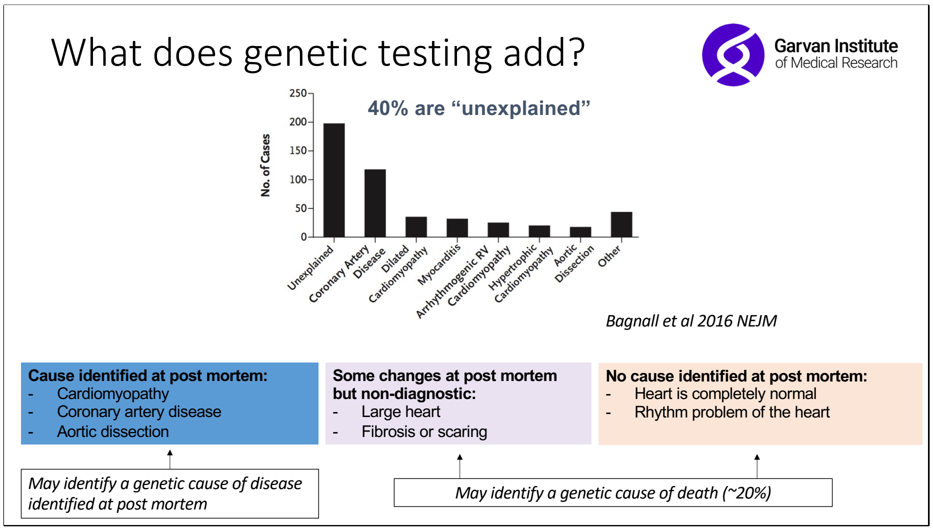


Slide outlines what genetic testing may add in each group.

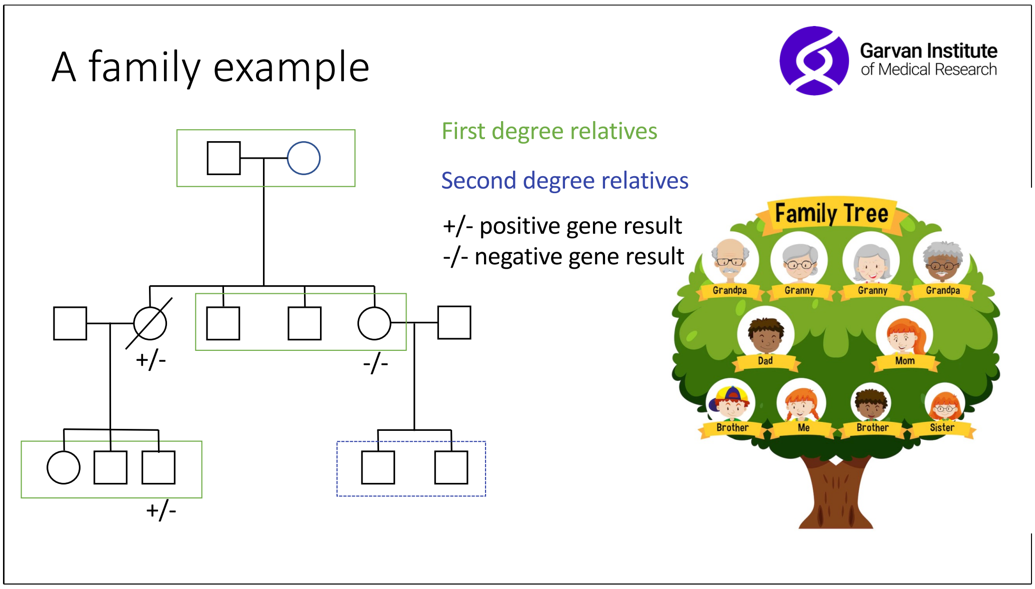


This slide is animated.
In this slide we explain what a family tree is, although they may be familiar with the tree on the right, the style on the left is what the team draw when we're investigating a young sudden death. General explanation of the symbols. We use this to explain who are first degree relatives, who are second degree relatives and highlight the utility of genetic testing in family members.

### What happens next?

- If a cause of death is **not** identified?
  - Repeat clinical screening?
  - Further genetic testing in the future?

Discussion around what happens if a cause of death is not identified.

### Making a plan for my family

Garvan Institute of Medical Research

- Who is in my family? (Draw your family history)
  - Anyone with:
    - heart disease?
    - rhythm problems? e.g., atrial fibrillation (AF)?
    - fainting/ blackouts?
    - SIDS?
    - epilepsy?
  - Who are the relatives in my family who need to be seen by a cardiologist?
  - How often should my relatives be screened by a cardiologist?

This slide aims to prompt participants on the sorts of questions or medical history they should consider when thinking about clinical screening in their family.

#### Preparing children for clinical screening

Garvan Institute of Medical Research

- Talk to your kids about the type of tests (ECG, echocardiogram)
- Explain what the tests involve in simple terms
  - ECG – dots on the chest, looks at rhythm of the heart
  - Echo – ultrasound of the heart, gel on chest and a special wand to look inside the body

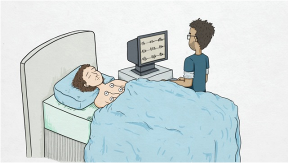

A slide to help those approach clinical screening in children. Children

#### Coping with uncertainty in family screening process?

Garvan Institute of Medical Research

- What helped for you when going through this process?

Open discussion on the process of clinical screening and genetic testing.

### Thank you for attending!

Garvan Institute
of Medical Research

- After today's session:
  - What are you going to do to look after yourself now?
  - If you feel distressed, who can you talk to?
    (family, friend, Beyond Blue – 1300 22 4636)
  - Please contact our team if you need further assistance
  - Any questions or anything you'd like to add?
  - Post session 2 survey will be emailed out to you

This slide is repeated at the end of each session as a prompt to consider how the participants are going to care for themselves now recognizing the discussion may have brought up some emotions for them.

#### Dates for your diary

Garvan Institute of Medical Research

- Session 3 – date and time
- Session 4 – date and time

**SESSION 3**

COPE-SCD: an online community supporting families after sudden cardiac death

Online session 3

Date

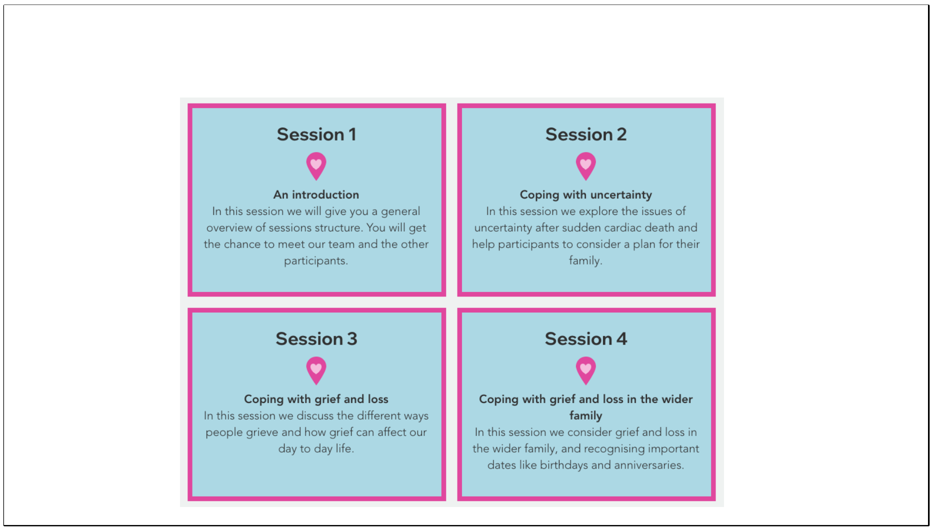


This slide will be repeated each session and just gives context on where today's session fits in and where we'll head next week.

#### Housekeeping

- **Confidentiality** – after the session if you're sharing with a friend about what was discussed, please don't use names
- **Selfcare** – Some of what you hear might trigger emotions, you can turn off your camera, go get some water, stand up etc.
- **Peer support** is a key part of this program – sharing experiences is helpful

LY

#### Checking in after session 2

- How did you go after last week?
- Any questions?

Regular check in after the previous week.

### Today

- We’re focusing on YOU!
- Next week, we’ll focus on your family and your community

### What does grief look like?

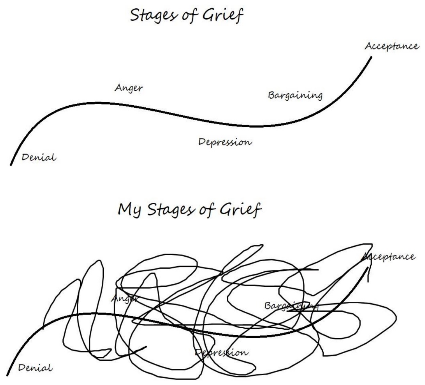


Source: https://www.facebook.com/AGriefTrajectory/photos/a.119153108736961/119823752003230/?type=3

When people think or talk about grief, they often imagine a scenario like the top of this picture.
However, in reality, grief often looks like the picture at the bottom. Messy, backwards and forwards etc.
Lots of literature de-bunking the Kubler-Ross five stages of grief but this picture is to help group members to talk.
Open up to the group and ask if these pictures resonate with them?

### Grief

- Grief is a natural process
- Each person’s journey is unique
- Different members of your family and community will grieve differently at different times
- Grief after sudden cardiac death can complex
  - Sudden, loved one often young, loss can be traumatic, grappling with uncertainty (genetics, unknown cause of death, family screening)

General information about grief

### Common responses to grief

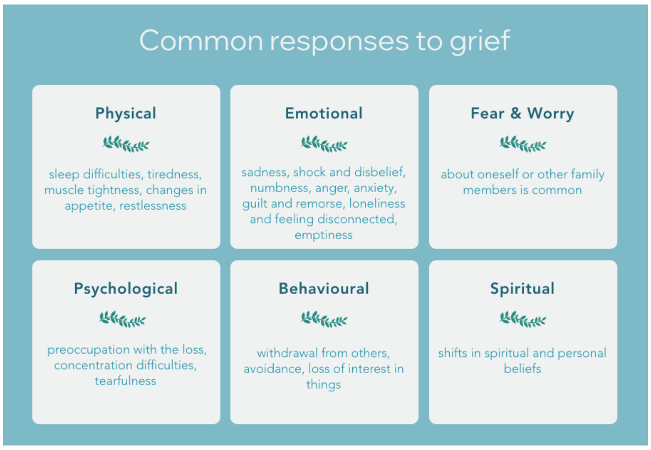


Secondary loss
Screenshot is taken from our website www.copescd.org.au

### Coping strategies

- What have you noticed that helps you?
- Good days vs bad days vs ok days/ moments
  - Routine:
    - eating regularly
    - sleep routine
    - gentle exercise
    - be alone or need company?
    - One thing a day
    - time in nature
    - looking after physical health
    - communicating your needs
    - allow others to support
  - Expectations of how grief should look
  - Self compassion

Talking points/ prompts for discussions.

### Thank you for attending!

- After today's session:
  - What are you going to do to look after yourself now?
  - If you feel distressed, who can you talk to?
    (family, friend, Beyond Blue – 1300 22 4636)
  - Please contact our team if you need further assistance
  - Any questions or anything you'd like to add?
  - Post session 3 survey will be emailed out to you

#### Dates for your diary

- Session 4 – date and time

#### SESSION 4

COPE-SCD: an online community supporting families after sudden cardiac death

Online session 4

Date

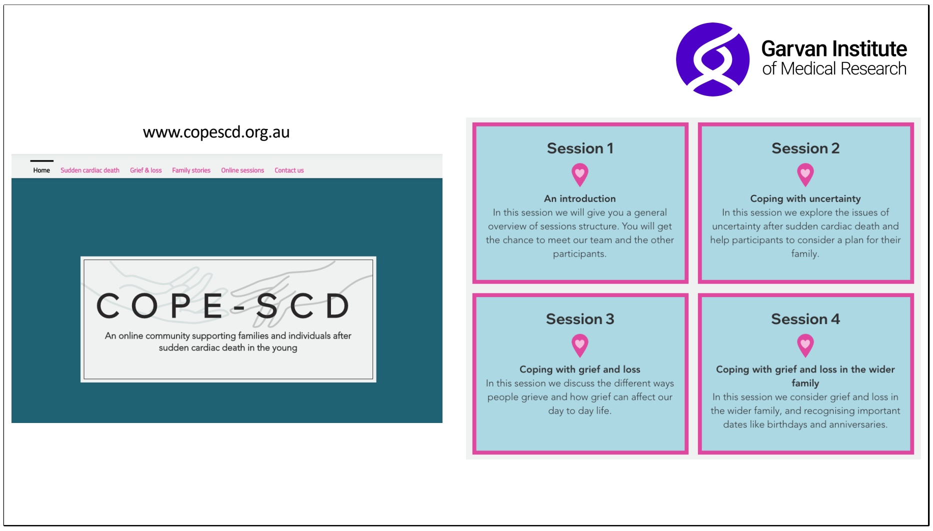


Reminder about the COPE-SCD program as a whole and more information available on our website.

### Checking in after session 3

- How did you go after last week?

### Today

- The goal today is to spend a bit of time on coping in the family/community context

### Family/ Community

Garvan Institute of Medical Research

- We recognise that "family" means different things to different people
- Today family means the community around you
- It may include:
  - DNA/ genetic family
  - Extended DNA family
  - Urban family (also known as friends)
  - Sporting family
  - Other important people around you

Description of "family"

### Your family and friends

Garvan Institute of Medical Research

- Family/friends may experience grief differently
- Some may prefer to share and others may need time alone
- How do people in your family/community grieve similarly/ differently?
- What have you found helpful in supporting each other (in your family/community)?

General discussion on grieving in the wider family.

### Your community

Garvan Institute of Medical Research

- What things do your family/friends do that is helpful/unhelpful?
- Past vs now
- Communication and boundaries

#### Anniversaries/ birthdays

Garvan Institute of Medical Research

- Anniversaries, birthdays and milestones are allowed to be hard - having a plan to care for oneself and each other around those times can help you.
- Each family/community may find their own unique way of honouring their loved one during one of these milestones
- A place you may visit
  - Journaling
  - Photos
  - Sharing of a favourite meal, activity, songs
- In what ways do you mark these milestones?

This slide is to give some prompts on approaching anniversaries/ birthdays or even missed milestones such as graduations.

### Reflections

Garvan Institute of Medical Research

- Thank you for being a part of the group and sharing
- Remind yourself grief is a natural process and allow yourself time to grieve individually and as a family
- Would like to share any final reflections/questions on the last 4 weeks?

A space for final reflections

### Thank you for attending!

Garvan Institute of Medical Research

- After today's session:
  - What are you going to do to look after yourself now?
  - If you feel distressed, who can you talk to?
    (family, friend, Beyond Blue – 1300 22 4636)
  - Please contact our team if you need further assistance
  - Any questions or anything you'd like to add?
  - Post session 4 survey will be emailed out to you

### Moving forward

Garvan Institute of Medical Research

- If you would like to connect with each other after this meeting, feel free to put your email address in the chat
- We are working with EndUCD.org who will have an ongoing support group – if you'd like to be kept in the loop about this please let Laura know
- There will be a couple of follow-up emails regarding the next steps in our evaluation
- A helpful website regarding children and grief
  - https://childhoodgrief.org.au/

Outlining methods for participants to connect outside of the program.

#### Still to come

Garvan Institute of Medical Research

- Post session 4 survey

- Final questionnaire
- Interview with Laura
- More details to come in your email

This slide has details about the session evaluations.

**Table S1** – Summary of post session surveys. P = participant

| Session | Median (range) | Percent helpful/ very helpful | Example quotes on what they liked most about each session |
|---|---|---|---|
| One (n=12) | 8 (7-10) | 83% | *“It was good to meet others in a similar situation and to hear about their stories - the similarities in particular, as it made you feel as though you're less alone”* P8 |
| Two (n=10) | 10 (8-10) | 100% | *“I found it very informative and addressed some of the big questions that family members face”* P4 |
| Three (n=10) | 9 (8-10) | 100% | *“Hearing about other people's firsthand experiences in coping with grief and being able to relate so well to those experiences”* P3 |
| Four (n=11) | 9 (5-10) | 82% | *“Sharing and listening to other’s experiences. Getting a few ideas of how to support myself better by being more direct/upfront”.* P6 |
